# Single-Channel Polysomnographic Signals Enable Accurate Age Estimation and Brain-Age Gap Biomarker Quantification

**DOI:** 10.1101/2025.08.07.25333233

**Authors:** Mattson Ogg, William G. Coon

## Abstract

Biological age estimation based on physiological signatures such as brain activity has emerged as a valuable biomarker; the gap between an individual’s predicted biological age and their chronological age is increasingly being linked to diverse health and cognitive outcomes. This study explores polysomnographic (PSG) recordings of sleep to evaluate how well diverse physiological signals—including EEG, ECG, EOG, and EMG—can support accurate age estimation, both individually and in combination. PSG serves as an ideal platform for age estimation due to its standardized data collection protocols, abundant public data resources, and its capture of well-documented age-related changes in sleep architecture that also relate to chronic health outcomes. Accordingly, we trained transformer-based neural network models on over 10,000 nights of public PSG data and performed rigorous internal and external validation. The best models achieved age estimates with an absolute error of 5-10 years on held out data from a single channel of electroencephalography (EEG). Interestingly, accuracy in these models was stage-dependent, with higher concentrations of N2 sleep in a given input sequence yielding more precise estimates than sequences dominated by other sleep stages. Age overestimation was associated with worse depression and cognitive performance outcomes. Electrocardiogram (ECG) signals, although less accurate overall, tended to overestimate age in association with health conditions such as elevated blood pressure, higher body mass index, and sleep apnea. Despite strong performance, generalization to external data remains a challenge (age estimation errors increase between internal validation and external data by at least 3 to 5 years). These findings show that non-invasive sleep-derived electrophysiological signals, particularly EEG, can support age estimation with accuracy comparable to functional MRI-based (5 to 11 years estimation error), yet at a fraction of the cost. The proliferation of consumer sleep monitoring devices, coupled with these highly accurate electrophysiological models, makes population-scale, at-home assessment of biological brain aging increasingly feasible.^1^

## Introduction

Aging is a complex multisystem process that can indicate health information about numerous organs and bodily systems. Advanced aging increases risk of disease within multiple physiological systems (Kaeberlein et al., 2015) and many common neurodegenerative disorders have also been specifically associated with age-related biological processes (Hou et al., 2019). It will thus be critical to understand healthy and unhealthy aging profiles as we develop and evaluate interventions aimed at slowing biological aging (Foley et al., 2004; Ibrahim et al., 2024; Kaeberlein et al., 2015), extending the healthspan, or preserving cognitive function over the lifespan (Carmona & Michan, 2016; Spira et al., 2014). Modern increases in life expectancy, which place growing segments of the population at risk for age-related diseases and neurodegenerative disorders (Dorsey et al., 2013; Feigin et al., 2020; García-Perdomo et al., 2019), highlight the urgent need for interventions that can support healthy aging and to gain a deeper understanding of the biology of senescence. Indeed, neurological and psychiatric disorders, which become more prevalent with age, are now a leading cause of disability (Feigin et al., 2020; Foley et al., 2004; Van Schependom & D’haeseleer, 2023). Hence, brain health biomarkers are increasingly of interest.

Brain structure and function have supported the best estimates of an individual’s age relative to other physiological systems (Tian et al., 2023). Thus, one promising biomarker to quantify brain health is the difference between an individual’s estimated age (as derived from their brain structure or function, sometimes called “brain age”), and their chronological age, often referred to as the “brain-age gap” (Cole & Franke, 2017; Franke & Gaser, 2019). A larger brain-age gap, indicative of advanced biological aging of the brain, has been observed in multiple clinical populations compared to age-matched controls, and the gap often widens in the presence of more advanced disease (Franke & Gaser, 2019; Kaufmann et al., 2019; Ogg & Kitchell, 2025). The utility of the brain-age gap as a biomarker stems from the fact that many neurological disorders share aging-related mechanisms, leading to accelerated structural or functional changes that can be detected by age estimation models (Cole & Franke, 2017).

A majority of the work on brain-age estimation and the development of brain-age gap biomarkers has been carried out using structural MRI, particularly T1-weighted scans (Cole & Franke, 2017; Franke & Gaser, 2019). The availability of large datasets, such as the UK Biobank (Miller et al., 2016), has enabled the development of highly accurate models that predict age with a mean absolute error (MAE) of approximately four years (Tian et al., 2023), using features such as gray matter volumes and cortical thickness derived from healthy participants. Leveraging simple 3D convolutional neural networks that learn features directly from raw T1-weighted images has further reduced age-estimation error to as low as 2.14 years MAE (Peng et al., 2021). These methods have revealed significant associations between brain-age gaps and neuropsychiatric disorders, particularly Alzheimer’s disease (AD) and mild cognitive impairment (MCI; Lee et al., 2022; Leonardsen et al., 2022; Millar et al., 2023; Ogg & Kitchell, 2025; Tian et al., 2023). Similarly, brain-age gap analyses have been extended to other conditions, such as schizophrenia, multiple sclerosis, bipolar disorder (Leonardsen et al., 2022), and traumatic brain injury (Cole et al., 2015; Spitz et al., 2022). Functional MRI (fMRI) has also been used to estimate age, albeit with reduced accuracy (5 to 11 years of age estimate error) relative to structural images, and with similar associations found between Alzheimer’s disease and age-overestimation (Bi et al., 2024; Millar et al., 2022; Ogg & Kitchell, 2025; Tian et al., 2023).

These results must be interpreted with a certain measure of caution, as estimation errors typically increase when models are applied to new datasets or populations that were not included in the model training phase (Leonardsen et al., 2022). Dufumier and colleagues (2022) recommend addressing this issue in two stages of validation: internal validation using held out data from the same study population used for training the model, and external validation using data from entirely new cohorts or imaging sites. External validation is crucial for assessing the generalizability of models, particularly when they have been trained on limited data or datasets from only one or a few imaging centers.

Despite its strengths, MRI-based brain-age estimation has notable limitations. MRI is expensive, inaccessible to some individuals (e.g., those with metal implants or claustrophobia), and not portable. These shortcomings for MRI also complicate the characterization of higher density brain-age sampling, for example from a large cohort of participants across multiple days. Moreover, the predominant use of structural T1-weighted images ignores features of brain function that might change over the lifespan (though these are captured in some age estimation work using fMRI, Millar et al., 2022, 2023; Ogg & Kitchell, 2025). As a more accessible alternative, some studies have explored brain-age estimation using electrophysiological signals such as electroencephalography (EEG). Engemann and collaborators (2022) achieved MAEs in the range of 6.5 to 8 years via cross-validation using several benchmark EEG datasets. Banville and team (2024) went further, collecting a large at-home dataset via a commercial EEG device and estimating age by cross-validating among individuals, achieving an MAE of 9.24. Moreover, age estimation could be improved with this device when using EEG data collected during sleep (MAE = 7.57), finding wake and rapid eye movement (REM) stages produce worse age estimates than non-REM stages N2 and N3, which were more accurate.

Sleep, often studied using EEG along with other physiological signals in a polysomnography (PSG) record (Silber et al. 2007; Rechtschaffen & Kales 1968), offers a particularly promising platform for assessing biological age. This is in large part due to the multi-system nature of its standard assessment (i.e., PSG includes measures of brain activity, cardiopulmonary function, respiration, eye motility, etc.; Keenan & Hirshkowitz, 2011), the large volume of publicly available sleep EEG datasets (G.-Q. Zhang et al., 2018), and the well-characterized age-related changes in sleep architecture observable via EEG (Li et al., 2018; Mander et al., 2017; Ohayon et al., 2004). These sleep changes are also accelerated in age-related neurodegenerative diseases such as Alzheimer’s and Parkinson’s (Ibrahim et al., 2024; Mander et al., 2017; Spira et al., 2014), as well as in other cardiovascular and respiratory pathologies (McNicholas et al., 2019; Wolk et al., 2005; Zee & Turek, 2006). Publicly available repositories of PSG data have already reached volumes (G.-Q. Zhang et al., 2018) substantial enough to support the development of advanced deep learning models for analyzing sleep physiology (Fiorillo et al., 2019; Hanna & Flöel, 2023; Perez-Pozuelo et al., 2020; Perslev et al., 2021).

Multiple groups have begun leveraging these data to develop models that estimate biological age from sleep. Sun and colleagues (2019) used spectral and temporal features extracted from EEG signals in sleep data to estimate age with an MAE of 7.6 years, finding that age-overestimation was associated with neurological and psychiatric conditions as well as diabetes and hypertension relative to healthy controls, thus providing evidence for an EEG-based brain-age gap that encodes clinically useful information. Ye and others (2020) extended these results to the prediction of dementia and all-cause mortality (Paixao et al., 2020). Brink-Kjaer and colleagues (2022) developed a neural-network based model to estimate age with errors as low as 5.8 years using EEG and other physiological signals derived from a PSG record. Age-gap estimates derived from these model outputs could be used to predict all-cause mortality and life expectancy. Zhang et al., (2024) obtained similar results (MAE = 6.18) using novel objective functions to train a transformer-based model architecture. Finally, age can be estimated accurately using models trained using self-supervised objectives that learn the structure of sleep on their own (Banville et al., 2021; Coon & Ogg, 2025; Ogg & Coon, 2024; Thapa et al., 2024), indicating that age-related changes are a clearly discernible feature in the latent structure of sleep that is interpretable by deep-learning methods.

Deep learning models have only recently been applied to age estimation from PSG signals. While initial results are promising, significant gaps in knowledge and methodology remain. First, it is unclear how age estimates derived from the different physiological systems captured by a PSG record might relate to different clinical outcomes. Polysomnography comprises recordings of activity from multiple physiological systems (e.g., neural, cardiac etc.), each of which could relate to different health conditions. Second, more work is needed to understand how best to model physiological data recorded during sleep. Transformer models, which excel at time-series modeling, offer one excellent solution for modeling these data (Coon & Ogg, 2024) but are only just beginning to be applied to age estimation (D. Zhang et al., 2024). Increasing the accuracy of age estimation may also increase the statistical power and discriminative potential for brain-age gap analyses. Optimizing these model architectures for operation on more portable platforms will also be critical for integration with the growing consumer market of sleep monitoring products. Such consumer devices and wearables open the door for widespread use of sleep-based health diagnostics, which could in turn improve public health by lowering barriers to access (Perez-Pozuelo et al., 2020).

In summary, the compelling advantages of accessible, cost-effective, and portable tools for brain-age estimation (in contrast with MRI) has led to an exploration of EEG and PSG signals. Sleep-based brain-age assessment in particular holds promise as a scalable and comprehensive measure of health that can be integrated into research, clinical, and consumer-use settings (Banville et al., 2024; Perez-Pozuelo et al., 2020). The sheer amount of sleep data available, along with increasingly sophisticated deep learning techniques, should also support better generalization of results than boutique studies, bringing the potential widespread utility of the brain-age gap as a biomarker of brain health within reach for a substantially broader population than if limited to MRI techniques alone.

In this report, we present a thorough examination of the efficacy of individual physiological PSG signals for the task of estimating the age of individuals and for a brain-age gap biomarker using a novel transformer-based model architecture adapted from audio signal processing (Baevski et al., 2020; Hsu et al., 2021) to learn physiological time series data. We provide rigorous evaluation of our model before undertaking brain-age gap analyses, adopting community best practices from the long history of work using structural (Dufumier et al., 2022) and functional MRI (Millar et al., 2022, 2023) for estimating age from brain signals. The contributions of our model and analysis include: relying on only minimally processed single channel inputs, using matched data across physiological domains for a comparison of age estimation using different physiological signals, and independence from sleep stage labels in age estimation (i.e., raw signals can be used directly, without requiring laborious or noisy post-processing). We find that using this model, most input signals can provide performance comparable to other techniques on internal validation data, but that external generalization is more challenging. However, when using only a single EEG channel from the PSG record as input, this model can produce highly accurate age estimates on unseen data sets. Moreover, brain-age gap measures derived from the model’s outputs are associated with measures of mood and cognitive disorders. Exploratory analyses carried out using ECG (another measure of a critical physiological system), produced brain-age gap measures that were aligned with other important cardiovascular health outcomes, although overall performance on external data was reduced. Increasing accuracy using non-neural physiological signals like ECG could further lower the barrier to accessing this valuable brain health biomarker, as many popular health wearables already measure ECG (e.g., AppleWatch) and in a form factor even less burdensome than already minimally obtrusive EEG wearables.

## Methods

### Data and Preprocessing

Multiple large-scale public PSG datasets were assembled for training testing and evaluation, involving over 10,000 records. Data were obtained from the National Sleep Research Resource (G.-Q. Zhang et al., 2018) and the training, validation and testing partitions generated for this work are described in Table 1. Seven of these datasets (Blackwell et al., 2011; Chen et al., 2015; H. Lee et al., 2022; Quan et al., 1997; Redline et al., 2012; Spira et al., 2008; Young et al., 2009) were used for the model training partition, which comprised 10,244 PSG records from 7,089 individuals. Following Dufumier and colleagues (2022), we created two validation partitions. The first one was designated as the “internal” validation partition, and was made up of data from 768 participants semi-randomly selected to be held out from the seven datasets used for training (one record each), thus aligning closely with the demographics of the training data. A second held-out dataset was designated as the “external” validation partition and comprises a completely new dataset to evaluate generalization. We selected the in-laboratory HomePAP (Rosen et al., 2012) dataset for external evaluation, for comparability with prior work where it is often held out from training as an external benchmark dataset (Brink-Kjaer et al., 2022; D. Zhang et al., 2024).

**Table 1.**
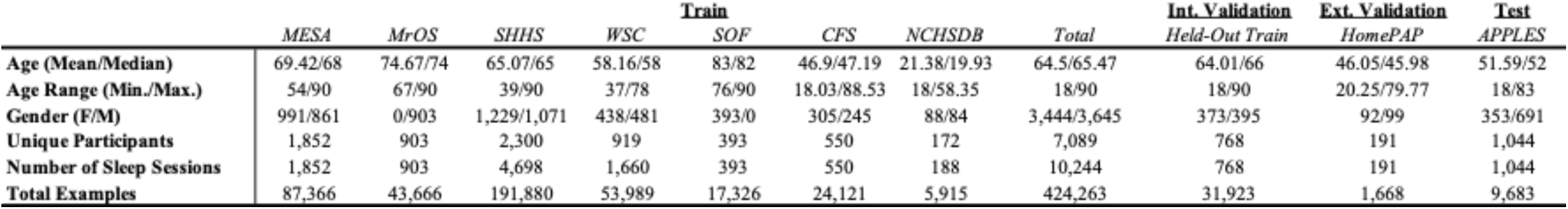
Data, Partitions and Demographics.

A third and final held-out dataset, the APPLES dataset (Kushida et al., 2012; Quan et al., 2011), was selected as a test dataset (also not used for training) since it contains extensive baseline physiological data as well as cognitive and mood testing, which made it a good candidate for brain-age gap analyses. We used data from Visit 3 (“DX”) which were released publicly (via the National Sleep Research Resource). We retained the following cognitive testing results and other metadata for use in brain-age gap analyses: Profile of Mood States (POMS), Epworth Sleepiness Scale (ESS), Sleep Apnea Quality of Life Index (SAQLI), Buschke Selective Reminding Test (BSRT), Pathfinder Task, Paced Auditory Serial Addition Task (PASAT), Psychomotor Vigilance Task (PVT) and the Sustained Working Memory Test measured at midday (SWMT; all from Visit 3), as well as Wechsler Abbreviated Scale of Intelligence IQ (WASI-IQ), Mini-Mental State Exam (MMSE), Beck Depression Inventory (BDI), Hamilton Rating Scale for Depression (HAM-D), Body Mass Index (BMI), weight and an apnea hypopnea index (AHI; all from baseline visit).

Because PSG records include dozens of signal sources, many of which are redundant, we selected a core subset of signals from each record to analyze. This minimal set included a single central EEG channel (C3 or C4), an EOG channel (left or right), ECG, EMG (“chin” where specified), and a piezoelectric respiration band recording (labelled “thorax” or “chest”). Each signal was then resampled to 100 Hz and robustly normalized (median-centered, scaled by the interquartile range, and clipped to +/- 20). Preprocessing was carried out using MNE-Python (Gramfort et al., 2013). We then extracted the data into 30-second epochs, retaining only those epochs that were assigned a sleep stage label (e.g., AASM Silber et al., 2007) in the accompanying sleep-scoring record. This was done to improve data quality for age estimation (but note that sleep stage labels were not used for model training as the model was trained on continuous time series data). Sleep stage labels were only later used for post-hoc analyses after the model was trained. Epochs were then arranged into sequences of 101 epochs (50.5 minutes of data) to serve as input to the model. This duration was chosen to balance maximum exposure to within-sleep-cycle sleep stage dynamics (for which longer sequences are preferred) against a maximum number of training examples within a single overnight (for which shorter sequences are preferred) and GPU memory limits. Data from the seven training datasets were arranged into sequences with a 25-epoch hop size to augment the size of our training corpus (approximately 4-times oversampling), and to expose the model to different views of a sleep cycle during training. The held-out data from the HomePAP and APPLES datasets were sequenced with no overlap because in this study they are used only for evaluation. We removed any PSG records or sequences with missing signals, to enable matched comparisons across PSG signal modalities. We also removed PSG records for participants under 18 years of age or if no age was listed. This was done to allow age estimation models to focus on features of adult sleep, which 1) exhibit marked differences from adolescent sleep and 2) might plausibly be related to brain health.

### Age Estimation Model Architecture and Training

We trained a neural network model to estimate the age of each participant (at the time of the corresponding sleep study) given a 50.5 minute physiological time series as input. For this we adapted a transformer architecture inspired by work on speech recognition (another field that learns features from continuous time series data; Baevski et al., 2020; Hsu et al., 2021), which has previously been shown to perform well analyzing sleep data (Coon & Ogg, 2024; Ogg & Coon, 2024) with multiple PSG signals (Coon et al., 2025; Coon & Ogg, 2025). The architecture, described in Figure 1 and Table 2, begins with feature extraction by first passing input through a series of seven one-dimensional convolutional layers that are each followed by a layer-norm and a Gaussian Error Linear Unit (GELU) activation function. Next, extracted features are linearly projected and positionally encoded before submission to a series of twelve transformer encoder layers. The convolutional layers learn local patterns in the data, while the transformer layers learn longer-term temporal patterns within each sequence (i.e., within and across 30-second sleep stage epochs in the input sequence). The output of the stack of transformer layers is averaged over time, linearly projected and passed through GELU activation functions, and finally passed to a single-unit output layer that produces an age estimate for the input sequence. A smaller version of this model was also trained to examine model performance as a function of model size, given results from ablation experiments in (Coon & Ogg, 2024) that showed robust performance from reduced-size model architectures (small model parameters described in Table 2).

**Figure 1.**
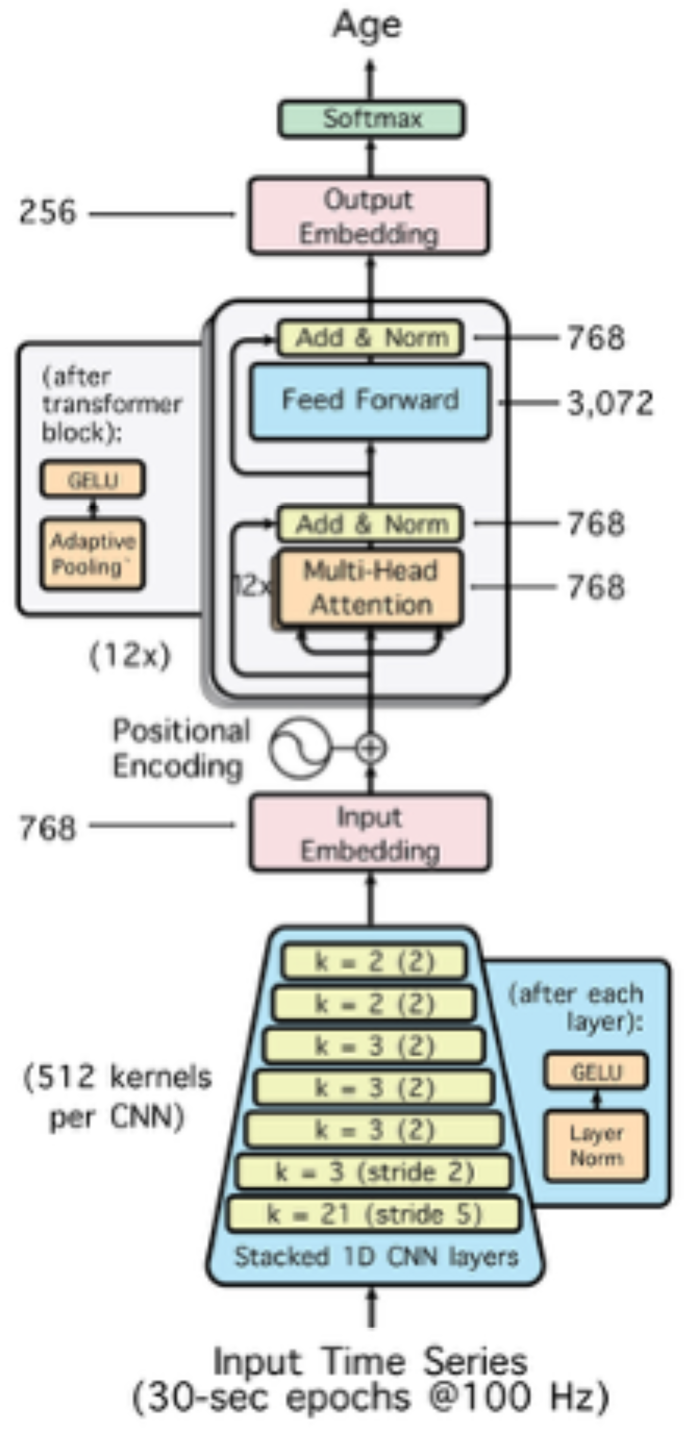
Schematic of Model Architecture. The convolutional layers form a front-end that first processes the time series data, learning a set of features (similar to filters) that are passed on to the transformer layers after projection and positional encoding. The transformer layers learn temporal patterns over the sequence of features output by the convolutional front-end. The convolutional layers process “micro” level structure in the sleep data (< 30-second epochs), while the transformers process “macro” level structure (> 30-second epochs) as well.

**Table 2.** Details of Model Architecture.

| Layer | Parameters | Large | Small |
| --- | --- | --- | --- |
| CNN Encoder | Strides | (5,2,2,2,2,2,2) |  |
|  | Kernel Width | (21,3,3,3,3,2,2) |  |
|  | Channels | 512 | 128 |
| Transformer | Layers | 12 | 2 |
|  | Embedding Dim | 768 | 384 |
|  | Inner FFN Dim | 3072 | 768 |
|  | Layerdrop Prob | 0.05 |  |
|  | Attention Heads | 12 | 6 |
| Average Pool 1D | n | 1 |  |
| Embedding | dim | 256 | 128 |
| Output | dim | 1 |  |
| Activation |  | gelu |  |
| Model Size | Trainable Params | 96.9 M | 3.9 M |
|  | MB | 403.2 | 23.4 |

A set of unimodal models was fit for each physiological signal input (EEG, EOG, EMG, ECG and respiration). Each unimodal model was trained to optimize a mean-squared error loss between the model output and the participant’s age at the visit corresponding to each 101-epoch input segment (with all age values in the loss scaled down by 100 for numerical stability during training). Each model was trained for 40 epochs with a peak learning rate of 0.0005. The learning rate ramped up linearly over the first 15 epochs from a starting factor that was 2% of the peak learning rate, and was then linearly ramped back down from the peak learning rate to 0.001% of the peak over the remaining epochs. Training used a batch size of 32 and an Adam optimizer.

### Model Evaluation

For each model, we saved the weights corresponding to the epoch that achieved the lowest loss on the internal validation partition for use on the external validation and testing data. Further evaluation was carried out by averaging the age estimates for each 101-epoch segment evaluated within a given PSG record to obtain a single age estimate for each night. We compared the session-level age estimates from each model with each participant’s ground truth chronological age using mean absolute error (MAE) and Pearson correlation. This was carried out for the internal validation set (of participants held out from the training dataset) and the external validation dataset (HomePAP). Following the recommendations in (Dufumier et al., 2022), the model with the lowest external validation MAE was used to evaluate the APPLES dataset and brain-age gap analyses.

### Brain-Age Gap Analysis

We evaluate the potential of a brain-age gap in our held out dataset using methods developed for functional data by Millar and colleagues (2022, 2023). Each clinical variable is entered into a linear model to estimate a brain-age gap variable derived from each model’s age estimates (specifically, estimated age minus chronological age) for each PSG record. These models also included chronological age, a variable for study site, a variable for race (with the largest classes set as the intercept), and a variable for sex (with the ‘male’ class set as the intercept) as predictors of no-interest to residualize brain-age delta and to offset potential biases in regression model performance (e.g., regression to mean, site-wise differences etc.). Note, a similar set of results was obtained when these predictors of no-interest were excluded. Thus, coefficients for the target clinical variables in these linear models can be interpreted as the amount of change on a given clinical scale per year of age overestimation by the model (holding other potential noise variables constant).

We use a similar method to evaluate which sleep stages contribute most to age estimation accuracy. Here, linear models are fit to the data at the sequence level (i.e., for each 101-epoch sequence evaluated prior to averaging by sleep session). Estimation error (specifically, the absolute value of the difference between model estimates and ground truth) was estimated as a function of five variables of counts for each sleep stage present in that sequence of data. Again, chronological age was included as a predictor of no-interest to offset age-wise estimation biases. Counts for each sleep stage were scaled down by a factor of four in the analysis of the internal validation partition to offset the oversampling. Counts for stage N1 were removed from these analyses due to low quantities of these epochs, poor inter-rater reliability for these stages (Danker-Hopfe et al., 2009; Rosenberg & Van Hout, 2013), and to help offset multicollinearity in these models (since otherwise for each sequence stage counts always summed to 101 across stages).

### Post-Hoc Exploratory Ablation and Combination Experiments

Additional exploratory analyses were undertaken to examine whether other hyperparameter choices might be useful for on-device deployment of our age estimation model architecture (since wearables with onboard processors are typically limited to small or compressed models only). Another age estimation model was trained for the best performing unimodal signal input using a much smaller instantiation of our transformer model (see Table 2). This was trained using the same hyperparameters and underwent the same evaluation as the full-size model.

Another exploratory analysis undertook the same brain-age gap analysis using cardiac activity captured by ECG. ECG signals are readily obtainable from widely available devices such as the AppleWatch. This accessibility could substantially reduce the barrier to estimating a brain-age gap compared to even the least intrusive sleep EEG wearables.

A final set of exploratory analyses sought to determine if age estimation performance could be optimized by combining signals that were input to an age estimation model. For these analyses a new set of transformer models was trained using the same procedures and hyperparameters as before, with the only modification being that these models took multiple (2 to 5) simultaneously recorded physiological signals as input: EEG+EOG (a common pairing for sleep stage classification; Coon & Ogg, 2024; Perslev et al., 2021), EEG+ECG (two disparate physiological signals with complementary clinical predictive utility), and a model that used all PSG signals (to see if it could learn a comprehensive view of brain age from PSG).

## Results

### Model Training and Validation

Polysomnography (PSG) data contains rich physiological signals that may support accurate age estimation across large public datasets and enable downstream analyses of brain-age gap biomarkers. In this study, we evaluated how effectively various standard PSG signals capture physiological features relevant to sleep-based modeling of age. To do this, we trained a set of five transformer models borrowing an architecture and hyperparameters previously shown to be effective for other time series modeling tasks (speech processing: Baevski et al., 2020; Hsu et al., 2021). Models were trained to estimate the age of individuals at each visit from parallel corpora of EEG, EOG, EMG, ECG, and respiratory data extracted from over ten thousand nights of sleep. Table 3 and Figure 2 summarize model performance on unseen participants from the training datasets.

**Figure 2.**
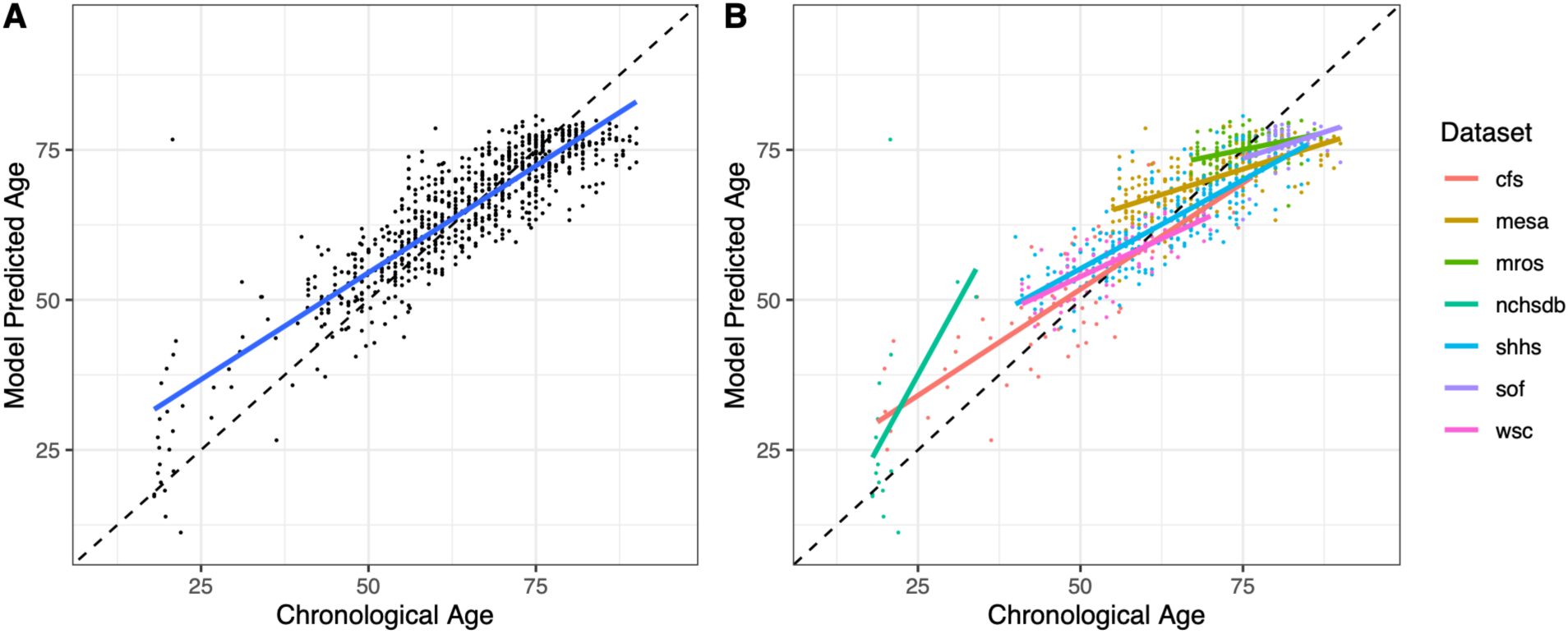
Performance of the unimodal EEG Model on the Internal Validation Partition. A) Overall performance and B) performance colored by dataset. Each point represents a single PSG record. Dotted lines indicate ideal performance, and solid lines indicate a linear fit to the data. See Table 3 for correlation statistics.

**Table 3.** Model Performance Across Datasets and Evaluation Partitions.

|  |  | Internal Validation |  |  |  |  |  |  | Ext. Val. | Test |  |
| --- | --- | --- | --- | --- | --- | --- | --- | --- | --- | --- | --- |
|  |  | MESA | MrOS | SHHS | WSC | SOF | CFS | NCHSDB | All | HomePAP | Apples |
| Uni-Modal Models | EEG | 5.71 / 0.66* | 3.37 / 0.36* | 4.74 / 0.81* | 4.06 / 0.75* | 5.8 / 0.4* | 7.25 / 0.85* | 11.12 / 0.52* | 5.15 / 0.88* | 10.18 / 0.66* | 8.07 / 0.72* |
|  | EOG | 6.36 / 0.64* | 3.43 / 0.47* | 4.93 / 0.8* | 5.15 / 0.59* | 4.55 / 0.22 | 7.93 / 0.8* | 10.21 / 0.51 | 5.46 / 0.88* | 10.43 / 0.52* | - |
|  | ECG | 6.81 / 0.43* | 4.64 / 0.28* | 7.08 / 0.5* | 7.11 / 0.36* | 5.8 / 0.19 | 9.73 / 0.66* | 14.77 / 0.23 | 6.98 / 0.78* | 16.69 / 0.16* | - |
|  | EMG | 7.27 / 0.29* | 3.9 / 0.38* | 7.76 / 0.36* | 6.08 / 0.21 | 4.4 / 0.26 | 9.41 / 0.69* | 20.78 / 0.39 | 7.09 / 0.77* | 14.24 / 0.24* | - |
|  | AIR | 7.14 / 0.45* | 3.64 / 0.27* | 7.66 / 0.42* | 5.39 / 0.4* | 3 / -0.21 | 9.81 / 0.64* | 5.11 / 0.51 | 6.51 / 0.82* | 15.01 / 0.18* | - |
| Post-Hoc Experiments | EEG+EOG | 5.22 / 0.74* | 3.59 / 0.49* | 4.15 / 0.86* | 3.89 / 0.78* | 4.18 / 0.27 | 5.56 / 0.91* | 7.33 / 0.33 | 4.51 / 0.92* | 10.74 / 0.69* | 9.91 / 0.44* |
|  | EEG+ECG | 6.63 / 0.45* | 4.05 / 0.4* | 5.67 / 0.69* | 4.88 / 0.52* | 5.49 / 0.35* | 8.11 / 0.79* | 10.79 / 0.21 | 5.9 / 0.84* | 14.36 / 0.35* | 10.96 / 0.44* |
|  | All Signals | 5.89 / 0.65* | 3.44 / 0.4* | 4.83 / 0.78* | 5 / 0.56* | 2.89 / 0 | 6.97 / 0.84* | 3.03 / 0.21 | 4.95 / 0.9* | 11.84 / 0.21* | 15.22 / 0.26* |
|  | EEG (Small) | 5.96 / 0.64* | 3.53 / 0.34* | 4.87 / 0.81* | 4.29 / 0.71* | 5.48 / 0.26 | 7.03 / 0.85* | 11.65 / 0.48 | 5.27 / 0.88* | 11.21 / 0.64* | 7.41 / 0.71* |

\* indicates correlation tests that survived FDR-correction

The unimodal EEG model (i.e., the model using only EEG as input) performed best on the internal validation dataset (MAE = 5.12 and *r* = 0.88, *p* < 0.001, overall), but most PSG signals could support reasonable overall performance (e.g., less than 7 years MAE) on these internal validation data. Internal validation presents the model with unseen participants, but still involves recording conditions and a distribution of targets that is closely matched to the training set (similar to cross-validation experiments), both of which likely result in an optimistic view of model performance. A stronger test of how a model might perform on new data comes from testing on completely new sets of data.

We evaluated each unimodal model on an additional external validation partition, comprising the HomePAP dataset, which is often used as a held out benchmark corpus for age estimation (Brink-Kjaer et al., 2022; D. Zhang et al., 2024). Again, the unimodal EEG model produced the best performance (MAE = 10.16, *r* = 0.66, *p* < 0.001; see Figure 3), despite study participants in the HomePAP data set being almost two decades younger than our training corpus on average. However, performance was considerably worse on the HomePAP data for non-EEG models. Due to EEG providing the best estimates of age on the HomePAP dataset compared to other single-signal models, we focus on the unimodal EEG model for subsequent analyses.

**Figure 3.**
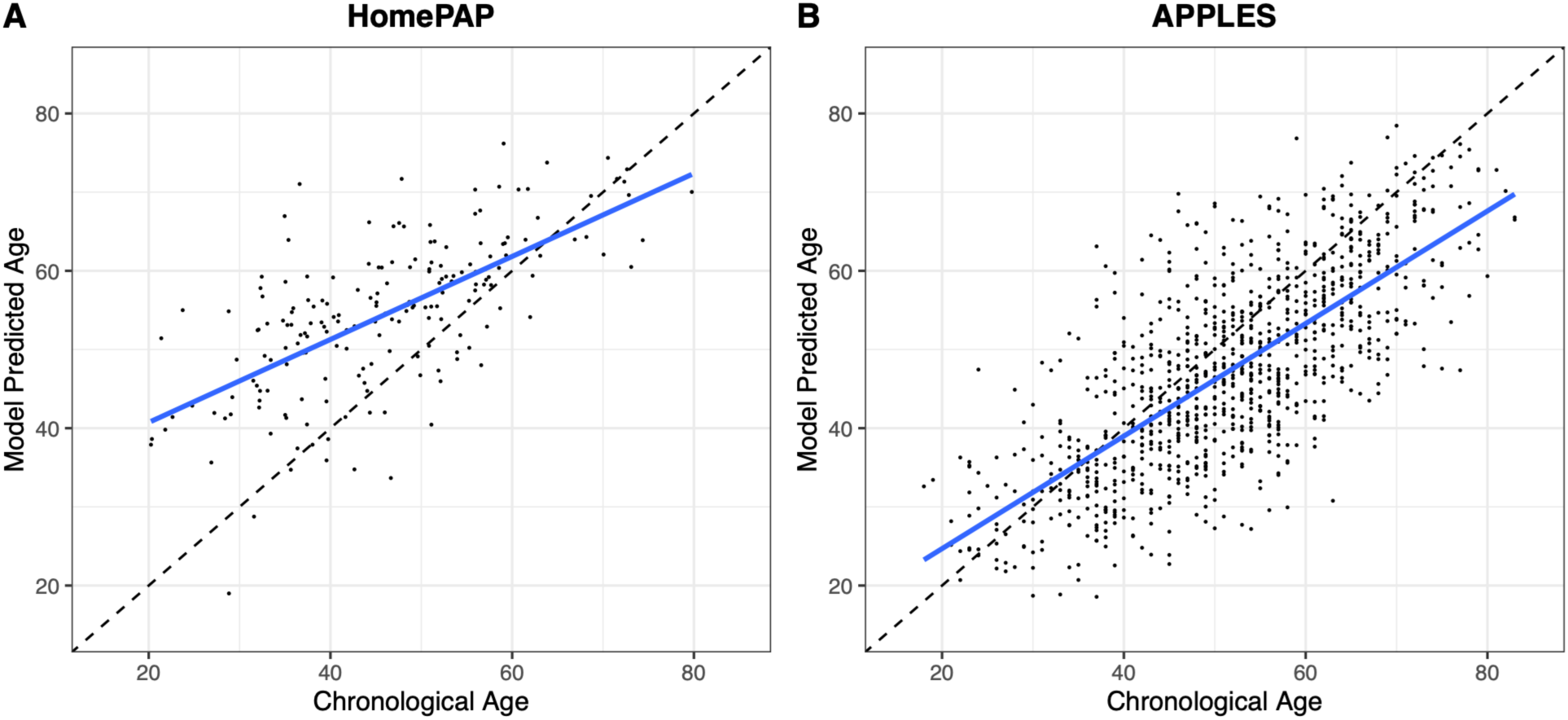
Performance of the unimodal EEG Model on the External Validation (HomePAP) and Testing (APPLES) Partitions. A) Model performance on the external validation, HomePAP dataset and B) model performance on the test, APPLES, dataset. Each point represents a single PSG record. Dotted lines indicate ideal performance, and solid lines indicate a linear fit to the data. See Table 3 for correlation statistics.

### Brain-Age Gap Assessment

EEG signals supported the most accurate brain-age estimation performance on an external dataset. Using this high-performing model, we wanted to assess the clinical utility of these age estimate outputs in the context of a brain-age gap analysis. For this, we evaluated an additional external corpus (APPLES) comprising over 1000 nights of sleep with extensive medical and cognitive testing data for each individual. These additional metadata could be used to compare the model’s estimation errors to clinical and cognitive outcomes. The hypothesis was that participants who present as being older might have additional unfavorable health or behavioral outcomes associated with an accelerated age phenotype.

The unimodal EEG model performed better on the APPLES dataset than on the HomePAP dataset, achieving a better MAE and higher correlation between model outputs and ground truth (MAE = 8.07 and *r* = 0.72, *p* < 0.001). Model outputs were used to generate a brain-age delta score (model age estimate - chronological age) for each sleep session, which we evaluated using the set of clinical metadata available in this dataset. Accelerated age estimates from the EEG model (positive brain-age delta scores) were associated with reduced Paced Auditory Serial Addition task performance (*b* = -0.018, S.E. = 0.006, *t* = -2.83, *p* < 0.01), higher weight (*b* = 0.012, S.E. = 0.006, *t* = 2.05, *p* < 0.05), worse Beck Depression Inventory scores (*b* = 0.179, S.E. = 0.051, *t* = 3.52, *p* < 0.001), and worse Hamilton Rating Scale for Depression scores (*b* = 0.166, S.E. = 0.066, *t* = 2.49, *p* < 0.05). We also observed marginally worse Sleep Apnea Quality of Life Index ratings (*b* = -0.126, S.E. = 0.070, *t* = -1.78, *p* = 0.075) and apnea hypopnea index scores (*b* = 0.0196, S.E. = 0.010, *t* = 1.87, *p* = 0.06). Counterintuitively, we also observed slightly improved Pathfinder task performance (*b* = 0.101, S.E. = 0.044, *t* = 2.27, *p* < 0.05) with accelerated brain-age estimates from the EEG model. Among these, the Paced Auditory Serial Addition task and Beck Depression Inventory age gap associations survived a false discovery rate correction for multiple comparisons. Thus, brain-age gap scores derived from our EEG model were primarily related to worse cognitive and mood outcomes.

### Exploratory and Post-Hoc Analyses

Additional analyses were carried out to examine different model optimization approaches and other domains of clinical and health information.

#### Brain-Age Gap for Less Accurate but Critical Physiological Systems

EEG supported the best age estimation performance for our transformer model. Reducing age estimation errors is important because it increases sensitivity for downstream brain-age gap analyses. However, our rigorous age-estimation evaluation pipeline could overlook other potentially useful signals from other physiological domains that might have clinical utility. In particular, ECG is a valuable indicator of cardiovascular health. While ECG performance on external data was generally much worse than EEG for estimating participants’ age, we wanted to undertake an exploratory brain-age gap analysis using this signal to examine its utility in this domain. Note, however, that this lower performance means that these results should be interpreted with caution.

Overall, the ECG age estimation model achieved worse performance on APPLES than the EEG age estimation model (MAE = 14.55, *r* = 0.29, *p* < 0.001). However, accelerated age estimates from the ECG model (positive age delta scores) were associated with reduced Buschke Selective Reminding Test performance (*b* = -0.118, S.E. = 0.057, *t* = -2.06, *p* < 0.05), decreased Paced Auditory Serial Addition task performance (*b* = -0.056, S.E. = 0.012, *t* = -4.80, *p* < 0.001) higher weight (*b* = 0.036, S.E. = 0.010, *t* = 3.45, *p* < 0.001), lower Wechsler Abbreviated Scale of Intelligence IQ scores (*b* = -0.136, S.E. = 0.040, *t* = -3.37, *p* < 0.001), higher body mass index (*b* = 0.203, S.E. = 0.072, *t* = 2.81, *p* < 0.01), and higher apnea hypopnea index scores (*b* = 0.050, S.E. = 0.019, *t* = 2.59, *p* < 0.01). Counterintuitively, we again observed slightly improved Pathfinder task performance (*b* = 0.302, S.E. = 0.082, *t* = 3.67, *p* < 0.001) with accelerated brain-age estimates from the ECG model. Among these, the Paced Auditory Serial Addition, Wechsler Abbreviated Scale of Intelligence IQ, BMI, AHI, weight and Pathfinder brain-age gap associations survived a false discovery rate correction for multiple comparisons. In sum, while less accurate, ECG estimates of accelerated aging were found to be associated with numerous negative health outcomes, but often differed from the results observed for EEG. Taken together, this suggests that PSG signals from both neural and non-neural physiological sources encode information that can index brain-age gap scores.

#### Signal Combination

Given that the health outcomes associated with model-estimated accelerated aging from ECG input in many cases differed from EEG, we explored whether age models that took combined signals as input would improve or diminish performance. In most cases, we found that adding signals beyond EEG (i.e., concatenating the signals of interest and providing a two-channel input to the model) hurt model performance on external data, likely from a reduced capacity of the model to delineate between multi-modal inputs.

We trained a set of additional models that combined EEG and EOG (two signals that are often combined for improved automatic sleep stage classification; Coon & Ogg, 2024; Hanna & Flöel, 2023; Perslev et al., 2021), EEG and ECG (to try and capture both brain-age gap outcomes) and a model that took all five PSG signals as input. These results are summarized in Table 3. The addition of any signal to EEG impaired performance overall.

#### Model Size

The model architecture employed in most of our analyses was adapted from architectures originally developed for speech recognition. We demonstrated that these large models are highly effective for brain-age estimation tasks. However, deploying such large models on portable or wearable platforms may be challenging. Additionally, prior research suggests that large transformer models could be excessive for certain supervised sleep estimation tasks (Coon & Ogg, 2024), with smaller transformer architectures often achieving performance levels nearly equivalent to those of significantly larger models.

To examine this possibility, we trained a substantially smaller transformer model—characterized by reduced front-end filters and a smaller transformer block comprising only two layers (detailed specifications provided in Table 2)—using identical data and training parameters as the larger model. This compact model demonstrated only slightly reduced performance on our internal validation set and moderately decreased performance on the external HomePAP validation dataset. Interestingly, the smaller model showed improved performance on the APPLES dataset (see Table 3). Furthermore, this smaller model replicated the general pattern of findings observed in the brain-age gap analyses conducted on the APPLES dataset, although some effects were attenuated or did not reach statistical significance compared to those obtained using the larger, full-scale model. Brain-age gap effects were still significant for the Paced Auditory Serial Addition task (*b* = -0.018, S.E. = 0.007, *t* = -2.67, *p* < 0.01), Beck Depression Inventory (*b* = 0.159, S.E. = 0.056, *t* = 2.86, *p* < 0.01), and AHI (*b* = 0.023, S.E. = 0.011, *t* = 2.10, *p* < 0.05), but the Hamilton Rating Scale for Depression (*b* = 0.129, S.E. = 0.073, *t* = 1.78, *p* = 0.075) was only marginally significant and all other effects were non-significant. Paced Auditory Serial Addition task and Beck Depression Inventory age-gap associations were marginally significant after false discovery rate correction. Taken together, these results suggest that transformers could be substantially reduced in size, sacrificing only a small degradation in performance, paving the way for real-time or on-device deployment.

#### Associations with Sleep Stages

Sleep is characterized by a phasic progression through distinct stages, transitioning from wakefulness to deep sleep and subsequently into rapid eye movement (REM) sleep throughout the night. Certain neural features observed during specific sleep stages may carry more information regarding an individual’s age compared to others. For example, slow-wave amplitude, which is specifically confined to non-rapid eye movement (NREM) sleep, is known to decrease substantially with age. Other stage-dependent metrics may have varying prominence across all stages (rather than being limited to a subset of sleep stages), and metrics of overnight sleep composition are also known to change with increasing age, including decreased amounts of N3 and increased amounts of N2 (see Li et al., 2022 and Mander et al., 2017 for review).

To examine the contribution of different sleep stages to sleep-age estimation accuracy, we tallied the number of epochs scored for each stage within each sequence of 101-epochs we evaluated and entered them into a linear model as predictors of the full-size unimodal EEG model’s errors for each sequence. For our internal validation partition, this analysis indicated that sequences with more N2 (*b* = -0.014, S.E. = 0.004, *t* = -3.27, *p* < 0.005) supported more accurate age estimation (i.e., for each N2 epoch in the sequence, age estimation error decreased by 0.014), while records with more wake epochs (*b* = 0.015, S.E. = 0.004, *t* = 3.83, *p* < 0.001) achieved significantly worse age estimation (i.e., for each wake epoch in the sequence, age estimation error increased by 0.015 years). For our external validation partition (the HomePAP dataset), the only dependence on sleep stages was that wake epochs (*b* = 0.068, S.E. = 0.017, *t* = 4.08, *p* < 0.001) were related to significantly worse age estimates (i.e., for each wake epoch in the sequence, age estimation error increased by 0.068 years). For the APPLES dataset, sequences with more N2 (*b* = -0.045, S.E. = 0.006, *t* = -7.70, *p* < 0.001) and N3 (*b* = -0.074, S.E. = 0.010, *t* = -7.27, *p* < 0.001) supported more accurate age estimation (i.e., for each N2 and N3 epoch in the sequence, age estimation error decreased by 0.045 and 0.074 years, respectively).

Given the associations we observed between sleep stages and age-estimation accuracy, and that N3 is more prominent earlier in the night while REM is more prominent later, it is possible that age estimation errors might also vary over the course of a night. However, in both validation and held-out test datasets, we found that age-estimation error was not strongly correlated with when a given sequence was drawn from over the course of a night, and the sign of the correlation varied across datasets (Internal Validation: *rs* = 0.01, *p* = 0.25; HomePAP: *rs* = -0.07, *p* < 0.01; APPLES: *rs* = 0.06, *p* < 0.001). This may be due to the model having been trained on examples from across the entire night (both early and late periods). Since some of the most prominent age-related patterns in sleep EEG signals have been tied to N3 sleep (for example, decreasing number and amplitude of N3-defining slow waves across the lifespan), it may be possible to further optimize model performance by targeting certain times of the sleep period most likely to exhibit age-related changes.

It is important to note that while we used sleep stage labels for this analysis, sleep stage information is *not* used by the age estimation models described here (c.f., Banville et al., 2024; Sun et al., 2019; Ye et al., 2020; D. Zhang et al., 2024). Our models estimate age directly from sequences of physiological time series data. This is advantageous because it both increases parsimony and eliminates a potential source of noise, as sleep stage labels themselves are subjective and exhibit substantial variability across scorers (see Danker-Hopfe et al., 2009, Rosenberg & Van Hout, 2013, and van Gorp et al., 2022 for discussion).

## Discussion

Improving brain-age gap models and our understanding of biological aging will be critical as we develop treatments to slow the progression of aging, preserve functionality in old age, and ameliorate the negative consequences of aging (Carmona & Michan, 2016; Foley et al., 2004; Kaeberlein et al., 2015). The need for these advances is made more urgent by a progressive increase in lifespan and the overall increasing age of the global population (Dorsey et al., 2013; Feigin et al., 2020; García-Perdomo et al., 2019). To meet this need we leveraged low cost, multimodal physiological sleep data from multiple repositories of polysomnographic records. Sleep provides a useful pathway for understanding aging due both to its status as an accessible (and potentially nightly) measure of biological function with clear landmarks related to aging (Mander et al., 2017) as well as a rapidly developing consumer product space that could support deployment platforms for models and analytics, in turn increasing access to healthcare and public health monitoring (Perez-Pozuelo et al., 2020).

In this study we trained large transformer models to estimate age from different single-channel selections of physiological signals contained within a sleep PSG record. Among these, neural signals captured by EEG supported the most accurate age estimation. Discrepancies between the model’s age estimates and the participants’ true chronological age were associated with mood, cognitive and medical outcomes on independent, held out datasets. While our model’s performance on some benchmark data sets is lower than other recent reports (Brink-Kjaer et al., 2022; D. Zhang et al., 2024), we restricted our models input primarily to single channels of physiological EEG data, and evaluate a much larger test corpus. This study remains one of a small few applying neural network models to time-varying physiological data for age estimation and is the only to comprehensively compare specific physiological signals. The EEG-only model achieved age estimation accuracy comparable to methods employing functional MRI (which is time-varying) and some structural MRI techniques (which are not time-varying), yet utilized overnight PSG studies, which are significantly more accessible and cost-effective than MRI.

A key contribution of this work is a comprehensive examination of the suitability of different peripheral physiological signals for accurate age estimation. The superior age-estimation accuracy of neural measures aligns with findings from recent EEG (Brink-Kjaer et al., 2022; Zhang et al., 2024) and MRI studies (Tian et al., 2023). Interestingly, like others (Brink-Kjaer et al., 2022), we observed that adding other signals alongside EEG generally led to less accurate age estimates. Finally, our exploratory analyses suggest there is some utility in querying multiple physiological systems (such as cardiovascular function), which could lead to complementary age-gap biomarkers and warrants further research.

Most signals we evaluated could support accurate age estimation within known populations or conditions, demonstrated by accurate performance from most signals on an internal validation set of held out subjects from the training corpora. In accordance with findings from age estimation using MRI, the performance of all models declined notably when evaluating external validation data from an independent study (Leonardsen et al., 2022; Peng et al., 2021). This underscores the importance of rigorous internal and external evaluation prior to brain-age gap assessments (Dufumier et al., 2022).

More work remains to identify which signal features drive model performance for age estimation. We began to explore this by examining the sleep stage composition of the PSG segments used for model training (50.5 mins of raw signal data). Similar to others (Banville et al., 2024; Zhang et al., 2024), we found that higher proportions of Wake within a segment increased estimation error, while higher proportions of N2 and N3 decreased error. This may be explained by the restriction of informative neural features—such as slow-wave amplitude and frequency—to sleep and to NREM sleep stages in particular. The observed decline in estimation accuracy with increased wakefulness within segments aligns with the understanding that sleep stages predominantly carry age-predictive information, consistent with established age-related physiological changes in sleep, including reduced slow-wave amplitude and number, decreased duration of stage N3, and increased duration of stage N2. These interpretations could be further confirmed using ablation techniques like SHapley Additive exPlanations (SHAP), which can identify input features that most strongly influence model output (see Brink-Kjaer et al., 2022 for an example)

There remains a vast design space for optimizing neural network architectures for sleep-based age estimation. In particular, improvements in training objectives, pretraining strategies, and transfer learning could significantly enhance model generalization and clinical applicability. Early work along these lines already shows promise (e.g., Banville et al., 2021; Coon & Ogg, 2025; Ogg & Coon, 2024; Thapa et al., 2026), suggesting that feature representations learned during age estimation may serve as powerful initializations for downstream clinical tasks. Additionally, improved methods for fusing information across different physiological modalities (e.g., EEG and ECG) represent a compelling direction for future research, especially given the mixed results we observed from signal combination in this study. Recent advances in cross-modal modeling and retrieval (Thapa et al., 2024) may offer a pathway forward.

Finally, our findings highlight a promising route for real-time, on-device age estimation using physiological sleep data. While large transformer models offer excellent performance, they can be computationally intensive and impractical for deployment on consumer-grade devices. Encouragingly, we found that a substantially smaller model—optimized with reduced parameters—achieved comparable performance in many settings and preserved several key brain-age gap associations. These results point toward the feasibility of running age estimation models directly on wearables or portable devices, lowering the barrier to personalized health monitoring at scale. As compute-efficient AI continues to evolve, such compact models may play a central role in enabling low-cost, privacy-preserving, and ubiquitous brain health assessment.

## Data Availability

Study used ONLY openly available human data hosted at the National Sleep Research Resource (NSRR) at sleepdata.org

https://sleepdata.org

https://osf.io/qw2d5

## Acknowledgements

We acknowledge support from the Independent Research and Development (IRAD) Fund from the Johns Hopkins Applied Physics Laboratory. While feedback from ChatGPT version 5.2 (hosted by OpenAI) was used to improve readability while preparing this report, all claims, analyses, and citations are the authors’ own.

## Footnotes

1 Model checkpoints along with training, evaluation and analysis scripts are available: https://osf.io/qw2d5

## Notes

### Competing Interest Statement

The authors have declared no competing interest.

### Funding Statement

This study was funded by an internal Research & Development grant by the Johns Hopkins Applied Physics Laboratory. No other funding, payment, or services were received in support of this work.

### Summary of Updates

We updated various parts of this manuscript.

## References

Baevski, A., Zhou, Y., Mohamed, A., & Auli, M. (2020). wav2vec 2.0: A Framework for Self-Supervised Learning of Speech Representations. Advances in Neural Information Processing Systems, 33, 12449–12460. https://proceedings.neurips.cc/paper/2020/hash/92d1e1eb1cd6f9fba3227870bb6d7f07-Abstract.html

Banville, H., Chehab, O., Hyvärinen, A., Engemann, D.-A., & Gramfort, A. (2021). Uncovering the structure of clinical EEG signals with self-supervised learning. Journal of Neural Engineering, 18(4), 046020. 10.1088/1741-2552/abca18

Banville, H., Jaoude, M. A., Wood, S. U. N., Aimone, C., Holst, S. C., Gramfort, A., & Engemann, D.-A. (2024). Do try this at home: Age prediction from sleep and meditation with large-scale low-cost mobile EEG. Imaging Neuroscience, 2, 1–15. 10.1162/imag_a_00189

Bi, S., Guan, Y., & Tian, L. (2024). Prediction of individual brain age using movie and resting-state fMRI. Cerebral Cortex, 34(1), bhad407. 10.1093/cercor/bhad407

Blackwell, T., Yaffe, K., Ancoli-Israel, S., Redline, S., Ensrud, K. E., Stefanick, M. L., Laffan, A., Stone, K. L., & for the Osteoporotic Fractures in Men (MrOS) Study Group. (2011). Association of Sleep Characteristics and Cognition in Older Community-Dwelling Men: The MrOS Sleep Study. Sleep, 34(10), 1347–1356. 10.5665/SLEEP.1276

Brink-Kjaer, A., Leary, E. B., Sun, H., Westover, M. B., Stone, K. L., Peppard, P. E., Lane, N. E., Cawthon, P. M., Redline, S., Jennum, P., Sorensen, H. B. D., & Mignot, E. (2022). Age estimation from sleep studies using deep learning predicts life expectancy. Npj Digital Medicine, 5(1), 1–10. 10.1038/s41746-022-00630-9

Carmona, J. J., & Michan, S. (2016). Biology of Healthy Aging and Longevity. Revista de Investigacion Clinica, 68(1), 7–16.

Carskadon, M. A., & Dement, W. C. (2011). Chapter 2 – Normal Human Sleep: An Overview.

Chen, X., Wang, R., Zee, P., Lutsey, P. L., Javaheri, S., Alcántara, C., Jackson, C. L., Williams, M. A., & Redline, S. (2015). Racial/Ethnic Differences in Sleep Disturbances: The Multi-Ethnic Study of Atherosclerosis (MESA). Sleep, 38(6), 877–888. 10.5665/sleep.4732

Cole, J. H., & Franke, K. (2017). Predicting Age Using Neuroimaging: Innovative Brain Ageing Biomarkers. Trends in Neurosciences, 40(12), 681–690. 10.1016/j.tins.2017.10.001

Cole, J. H., Leech, R., Sharp, D. J., & Initiative, for the A. D. N. (2015). Prediction of brain age suggests accelerated atrophy after traumatic brain injury. Annals of Neurology, 77(4), 571–581. 10.1002/ana.24367

Coon, W. G., Luna, D., Panagrahi, A., Reid, M., & Ogg, M. (2025). Getting More from Less: Transfer Learning Improves Sleep Stage Decoding Accuracy in Peripheral Wearable Devices (No. arXiv:2506.00730). arXiv. 10.48550/arXiv.2506.00730

Coon, W. G., & Ogg, M. (2024). Laying the Foundation: Modern Transformers for Gold-Standard Sleep Analysis and Beyond. 2024 46th Annual International Conference of the IEEE Engineering in Medicine and Biology Society (EMBC), 1–7. 10.1109/EMBC53108.2024.10782964

Coon, W. G., & Ogg, M. (2025). Foundation Models Reveal Untapped Health Information in Human Polysomnographic Sleep Data (p. 2025.07.15.25331562). medRxiv. 10.1101/2025.07.15.25331562

Danker-Hopfe, H., Anderer, P., Zeitlhofer, J., Boeck, M., Dorn, H., Gruber, G., Heller, E., Loretz, E., Moser, D., Parapatics, S., Saletu, B., Schmidt, A., & Dorffner, G. (2009). Interrater reliability for sleep scoring according to the Rechtschaffen & Kales and the new AASM standard. Journal of Sleep Research, 18(1), 74–84. 10.1111/j.1365-2869.2008.00700.x

Dorsey, E. R., George, B. P., Leff, B., & Willis, A. W. (2013). The coming crisis: Obtaining care for the growing burden of neurodegenerative conditions. Neurology, 80(21), 1989–1996. 10.1212/WNL.0b013e318293e2ce

Dufumier, B., Grigis, A., Victor, J., Ambroise, C., Frouin, V., & Duchesnay, E. (2022). OpenBHB: A Large-Scale Multi-Site Brain MRI Data-set for Age Prediction and Debiasing. NeuroImage, 263, 119637. 10.1016/j.neuroimage.2022.119637

Engemann, D. A., Mellot, A., Höchenberger, R., Banville, H., Sabbagh, D., Gemein, L., Ball, T., & Gramfort, A. (2022). A reusable benchmark of brain-age prediction from M/EEG resting-state signals. NeuroImage, 262, 119521. 10.1016/j.neuroimage.2022.119521

Feigin, V. L., Vos, T., Nichols, E., Owolabi, M. O., Carroll, W. M., Dichgans, M., Deuschl, G., Parmar, P., Brainin, M., & Murray, C. (2020). The global burden of neurological disorders: Translating evidence into policy. The Lancet Neurology, 19(3), 255–265. 10.1016/S1474-4422(19)30411-9

Fiorillo, L., Puiatti, A., Papandrea, M., Ratti, P.-L., Favaro, P., Roth, C., Bargiotas, P., Bassetti, C. L., & Faraci, F. D. (2019). Automated sleep scoring: A review of the latest approaches. Sleep Medicine Reviews, 48, 101204. 10.1016/j.smrv.2019.07.007

Foley, D., Ancoli-Israel, S., Britz, P., & Walsh, J. (2004). Sleep disturbances and chronic disease in older adults: Results of the 2003 National Sleep Foundation Sleep in America Survey. Journal of Psychosomatic Research, 56(5), 497–502. 10.1016/j.jpsychores.2004.02.010

Fox, B., Jiang, J., Wickramaratne, S., Kovatch, P., Suarez-Farinas, M., Shah, N. A., Parekh, A., & Nadkarni, G. N. (2025). A foundational transformer leveraging full night, multichannel sleep study data accurately classifies sleep stages. *Sleep*, zsaf061. 10.1093/sleep/zsaf061

Franke, K., & Gaser, C. (2019). Ten Years of BrainAGE as a Neuroimaging Biomarker of Brain Aging: What Insights Have We Gained? Frontiers in Neurology, 10. 10.3389/fneur.2019.00789

García-Perdomo, H. A., Zapata-Copete, J., & Rojas-Cerón, C. A. (2019). Sleep duration and risk of all-cause mortality: A systematic review and meta-analysis. Epidemiology and Psychiatric Sciences, 28(5), 578–588. 10.1017/S2045796018000379

Gramfort, A., Luessi, M., Larson, E., Engemann, D. A., Strohmeier, D., Brodbeck, C., Goj, R., Jas, M., Brooks, T., Parkkonen, L., & Hämäläinen, M. (2013). MEG and EEG data analysis with MNE-Python. Frontiers in Neuroscience, 7. 10.3389/fnins.2013.00267

Hanna, J., & Flöel, A. (2023). An accessible and versatile deep learning-based sleep stage classifier. Frontiers in Neuroinformatics, 17. 10.3389/fninf.2023.1086634

Hou, Y., Dan, X., Babbar, M., Wei, Y., Hasselbalch, S. G., Croteau, D. L., & Bohr, V. A. (2019). Ageing as a risk factor for neurodegenerative disease. Nature Reviews Neurology, 15(10), 565–581. 10.1038/s41582-019-0244-7

Hsu, W.-N., Bolte, B., Tsai, Y.-H. H., Lakhotia, K., Salakhutdinov, R., & Mohamed, A. (2021). HuBERT: Self-Supervised Speech Representation Learning by Masked Prediction of Hidden Units. *IEEE/ACM Transactions on Audio*, Speech, and Language Processing, 29, 3451–3460. 10.1109/TASLP.2021.3122291

Ibrahim, A., Cesari, M., Heidbreder, A., Defrancesco, M., Brandauer, E., Seppi, K., Kiechl, S., Högl, B., & Stefani, A. (2024). Sleep features and long-term incident neurodegeneration: A polysomnographic study. Sleep, 47(3), zsad304. 10.1093/sleep/zsad304

Kaeberlein, M., Rabinovitch, P. S., & Martin, G. M. (2015). Healthy aging: The ultimate preventative medicine. Science, 350(6265), 1191–1193. 10.1126/science.aad3267

Kaufmann, T., van der Meer, D., Doan, N. T., Schwarz, E., Lund, M. J., Agartz, I., Alnæs, D., Barch, D. M., Baur-Streubel, R., Bertolino, A., Bettella, F., Beyer, M. K., Bøen, E., Borgwardt, S., Brandt, C. L., Buitelaar, J., Celius, E. G., Cervenka, S., Conzelmann, A., … Westlye, L. T. (2019). Common brain disorders are associated with heritable patterns of apparent aging of the brain. Nature Neuroscience, 22(10), 1617–1623. 10.1038/s41593-019-0471-7

Keenan, S., & Hirshkowitz, M. (2011). Chapter 141 – Monitoring and Staging Human Sleep.

Kushida, C. A., Nichols, D. A., Holmes, T. H., Quan, S. F., Walsh, J. K., Gottlieb, D. J., Simon, R. D., Guilleminault, C., White, D. P., Goodwin, J. L., Schweitzer, P. K., Leary, E. B., Hyde, P. R., Hirshkowitz, M., Green, S., McEvoy, L. K., Chan, C., Gevins, A., Kay, G. G., … Dement, W. C. (2012). Effects of Continuous Positive Airway Pressure on Neurocognitive Function in Obstructive Sleep Apnea Patients: The Apnea Positive Pressure Long-term Efficacy Study (APPLES). Sleep, 35(12), 1593–1602. 10.5665/sleep.2226

Lee, H., Li, B., DeForte, S., Splaingard, M. L., Huang, Y., Chi, Y., & Linwood, S. L. (2022). A large collection of real-world pediatric sleep studies. Scientific Data, 9(1), 421. 10.1038/s41597-022-01545-6

Lee, J., Burkett, B. J., Min, H.-K., Senjem, M. L., Lundt, E. S., Botha, H., Graff-Radford, J., Barnard, L. R., Gunter, J. L., Schwarz, C. G., Kantarci, K., Knopman, D. S., Boeve, B. F., Lowe, V. J., Petersen, R. C., Jack, C. R., & Jones, D. T. (2022). Deep learning-based brain age prediction in normal aging and dementia. Nature Aging, 2(5), 412–424. 10.1038/s43587-022-00219-7

Leonardsen, E. H., Peng, H., Kaufmann, T., Agartz, I., Andreassen, O. A., Celius, E. G., Espeseth, T., Harbo, H. F., Høgestøl, E. A., Lange, A.-M. de, Marquand, A. F., Vidal-Piñeiro, D., Roe, J. M., Selbæk, G., Sørensen, Ø., Smith, S. M., Westlye, L. T., Wolfers, T., & Wang, Y. (2022). Deep neural networks learn general and clinically relevant representations of the ageing brain. NeuroImage, 256, 119210. 10.1016/j.neuroimage.2022.119210

Li, J., Vitiello, M. V., & Gooneratne, N. S. (2018). Sleep in Normal Aging. Sleep Medicine Clinics, 13(1), 1–11. 10.1016/j.jsmc.2017.09.001

Lundberg, S. M., & Lee, S.-I. (2017). A Unified Approach to Interpreting Model Predictions. Advances in Neural Information Processing Systems, 30. https://proceedings.neurips.cc/paper_files/paper/2017/hash/8a20a8621978632d76c43dfd28b67767-Abstract.html

Mander, B. A., Winer, J. R., & Walker, M. P. (2017). Sleep and Human Aging. Neuron, 94(1), 19–36. 10.1016/j.neuron.2017.02.004

McNicholas, W. T., Hansson, D., Schiza, S., & Grote, L. (2019). Sleep in chronic respiratory disease: COPD and hypoventilation disorders. European Respiratory Review, 28(153), 190064. 10.1183/16000617.0064-2019

Millar, P. R., Gordon, B. A., Luckett, P. H., Benzinger, T. L., Cruchaga, C., Fagan, A. M., Hassenstab, J. J., Perrin, R. J., Schindler, S. E., Allegri, R. F., Day, G. S., Farlow, M. R., Mori, H., Nübling, G., The Dominantly Inherited Alzheimer Network, Bateman, R. J., Morris, J. C., & Ances, B. M. (2023). Multimodal brain age estimates relate to Alzheimer disease biomarkers and cognition in early stages: A cross-sectional observational study. eLife, 12, e81869. 10.7554/eLife.81869

Millar, P. R., Luckett, P. H., Gordon, B. A., Benzinger, T. L. S., Schindler, S. E., Fagan, A. M., Cruchaga, C., Bateman, R. J., Allegri, R., Jucker, M., Lee, J.-H., Mori, H., Salloway, S. P., Yakushev, I., Morris, J. C., Ances, B. M., Adams, S., Allegri, R., Araki, A., … Xu, X. (2022). Predicting brain age from functional connectivity in symptomatic and preclinical Alzheimer disease. NeuroImage, 256, 119228. 10.1016/j.neuroimage.2022.119228

Miller, K. L., Alfaro-Almagro, F., Bangerter, N. K., Thomas, D. L., Yacoub, E., Xu, J., Bartsch, A. J., Jbabdi, S., Sotiropoulos, S. N., Andersson, J. L. R., Griffanti, L., Douaud, G., Okell, T. W., Weale, P., Dragonu, I., Garratt, S., Hudson, S., Collins, R., Jenkinson, M., … Smith, S. M. (2016). Multimodal population brain imaging in the UK Biobank prospective epidemiological study. Nature Neuroscience, 19(11), 1523–1536. 10.1038/nn.4393

Ogg, M., & Coon, W. G. (2024). Self-Supervised Transformer Model Training for a Sleep-EEG Foundation Model. 2024 46th Annual International Conference of the IEEE Engineering in Medicine and Biology Society (EMBC), 1–6. 10.1109/EMBC53108.2024.10782281

Ogg, M., & Kitchell, L. (2025). Optimizing Artificial Neural Network Models to Predict Brain-Age from Functional MRI (p. 2025.03.28.645964). bioRxiv. 10.1101/2025.03.28.645964

Ohayon, M. M., Carskadon, M. A., Guilleminault, C., & Vitiello, M. V. (2004). Meta-Analysis of Quantitative Sleep Parameters From Childhood to Old Age in Healthy Individuals: Developing Normative Sleep Values Across the Human Lifespan. Sleep, 27(7), 1255–1273. 10.1093/sleep/27.7.1255

Paixao, L., Sikka, P., Sun, H., Jain, A., Hogan, J., Thomas, R., & Westover, M. B. (2020). Excess brain age in the sleep electroencephalogram predicts reduced life expectancy. Neurobiology of Aging, 88, 150–155. 10.1016/j.neurobiolaging.2019.12.015

Peng, H., Gong, W., Beckmann, C. F., Vedaldi, A., & Smith, S. M. (2021). Accurate brain age prediction with lightweight deep neural networks. Medical Image Analysis, 68, 101871. 10.1016/j.media.2020.101871

Perez-Pozuelo, I., Zhai, B., Palotti, J., Mall, R., Aupetit, M., Garcia-Gomez, J. M., Taheri, S., Guan, Y., & Fernandez-Luque, L. (2020). The future of sleep health: A data-driven revolution in sleep science and medicine. Npj Digital Medicine, 3(1), 1–15. 10.1038/s41746-020-0244-4

Perslev, M., Darkner, S., Kempfner, L., Nikolic, M., Jennum, P. J., & Igel, C. (2021). U-Sleep: Resilient high-frequency sleep staging. Npj Digital Medicine, 4(1), 1–12. 10.1038/s41746-021-00440-5

Quan, S. F., Chan, C. S., Dement, W. C., Gevins, A., Goodwin, J. L., Gottlieb, D. J., Green, S., Guilleminault, C., Hirshkowitz, M., Hyde, P. R., Kay, G. G., Leary, E. B., Nichols, D. A., Schweitzer, P. K., Simon, R. D., Walsh, J. K., & Kushida, C. A. (2011). The association between obstructive sleep apnea and neurocognitive performance—The Apnea Positive Pressure Long-term Efficacy Study (APPLES). Sleep, 34(3), 303–314B. 10.1093/sleep/34.3.303

Quan, S. F., Howard, B. V., Iber, C., Kiley, J. P., Nieto, F. J., O’Connor, G. T., Rapoport, D. M., Redline, S., Robbins, J., Samet, J. M., & Wahl, ‡Patricia W. (1997). The Sleep Heart Health Study: Design, Rationale, and Methods. Sleep, 20(12), 1077–1085. 10.1093/sleep/20.12.1077

Redline, S., Tishler, P. V., Tosteson, T. D., Williamson, J., Kump, K., Browner, I., Ferrette, V., & Krejci, P. (2012). The familial aggregation of obstructive sleep apnea. American Journal of Respiratory and Critical Care Medicine. 10.1164/ajrccm.151.3.7881656

Rosen, C. L., Auckley, D., Benca, R., Foldvary-Schaefer, N., Iber, C., Kapur, V., Rueschman, M., Zee, P., & Redline, S. (2012). A Multisite Randomized Trial of Portable Sleep Studies and Positive Airway Pressure Autotitration Versus Laboratory-Based Polysomnography for the Diagnosis and Treatment of Obstructive Sleep Apnea: The HomePAP Study. Sleep, 35(6), 757–767. 10.5665/sleep.1870

Rosenberg, R. S., & Van Hout, S. (2013). The American Academy of Sleep Medicine Inter-scorer Reliability Program: Sleep Stage Scoring. Journal of Clinical Sleep Medicine : JCSM : Official Publication of the American Academy of Sleep Medicine, 9(1), 81–87. 10.5664/jcsm.2350

Silber, M. H., Ancoli, -Israel Sonia, Bonnet, M. H., Chokroverty, S., Grigg, -Damberger Madeleine M., Hirshkowitz, M., Kapen, S., Keenan, S. A., Kryger, M. H., Penzel, T., Pressman, M. R., & Iber, C. (2007). The Visual Scoring of Sleep in Adults. Journal of Clinical Sleep Medicine, 03(02), 121–131. 10.5664/jcsm.26814

Spira, A. P., Blackwell, T., Stone, K. L., Redline, S., Cauley, J. A., Ancoli-Israel, S., & Yaffe, K. (2008). Sleep-Disordered Breathing and Cognition in Older Women. Journal of the American Geriatrics Society, 56(1), 45–50. 10.1111/j.1532-5415.2007.01506.x

Spira, A. P., Chen-Edinboro, L. P., Wu, M. N., & Yaffe, K. (2014). Impact of sleep on the risk of cognitive decline and dementia. Current Opinion in Psychiatry, 27(6), 478–483. 10.1097/YCO.0000000000000106

Spitz, G., Hicks, A. J., Roberts, C., Rowe, C. C., & Ponsford, J. (2022). Brain age in chronic traumatic brain injury. NeuroImage: Clinical, 35, 103039. 10.1016/j.nicl.2022.103039

Sun, H., Paixao, L., Oliva, J. T., Goparaju, B., Carvalho, D. Z., van Leeuwen, K. G., Akeju, O., Thomas, R. J., Cash, S. S., Bianchi, M. T., & Westover, M. B. (2019). Brain age from the electroencephalogram of sleep. Neurobiology of Aging, 74, 112–120. 10.1016/j.neurobiolaging.2018.10.016

Thapa, R., He, B., Kjaer, M. R., Iv, H. M., Ganjoo, G., Mignot, E., & Zou, J. Y. (2024, March 1). SleepFM: Multi-modal Representation Learning for Sleep across ECG, EEG and Respiratory Signals. AAAI 2024 Spring Symposium on Clinical Foundation Models. https://openreview.net/forum?id=cDXtscWCKC

Thapa, R., Kjaer, M. R., He, B., Covert, I., Moore Iv, H., Hanif, U., Ganjoo, G., Westover, M. B., Jennum, P., Brink-Kjaer, A., Mignot, E., & Zou, J. (2026). A multimodal sleep foundation model for disease prediction. Nature Medicine. 10.1038/s41591-025-04133-4

Tian, Y. E., Cropley, V., Maier, A. B., Lautenschlager, N. T., Breakspear, M., & Zalesky, A. (2023). Heterogeneous aging across multiple organ systems and prediction of chronic disease and mortality. Nature Medicine, 29(5), 1221–1231. 10.1038/s41591-023-02296-6

van Gorp, H., Huijben, I. A. M., Fonseca, P., van Sloun, R. J. G., Overeem, S., & van Gilst, M. M. (2022). Certainty about uncertainty in sleep staging: A theoretical framework. Sleep, 45(8), zsac134. 10.1093/sleep/zsac134

Van Schependom, J., & D’haeseleer, M. (2023). Advances in Neurodegenerative Diseases. Journal of Clinical Medicine, 12(5), Article 5. 10.3390/jcm12051709

Wolk, R., Gami, A. S., Garcia-Touchard, A., & Somers, V. K. (2005). Sleep and Cardiovascular Disease. Current Problems in Cardiology, 30(12), 625–662. 10.1016/j.cpcardiol.2005.07.002

Ye, E., Sun, H., Leone, M. J., Paixao, L., Thomas, R. J., Lam, A. D., & Westover, M. B. (2020). Association of Sleep Electroencephalography-Based Brain Age Index With Dementia. JAMA Network Open, 3(9), e2017357. 10.1001/jamanetworkopen.2020.17357

Young, T., Palta, M., Dempsey, J., Peppard, P. E., Nieto, F. J., & Hla, K. M. (2009). Burden of sleep apnea: Rationale, design, and major findings of the Wisconsin Sleep Cohort study. WMJ: Official Publication of the State Medical Society of Wisconsin, 108(5), 246–249.

Zee, P. C., & Turek, F. W. (2006). Sleep and Health: Everywhere and in Both Directions. Archives of Internal Medicine, 166(16), 1686–1688. 10.1001/archinte.166.16.1686

Zhang, D., She, Y., Sun, J., Cui, Y., Yang, X., Zeng, X., & Qin, W. (2024). Brain Age Estimation from Overnight Sleep Electroencephalography with Multi-Flow Sequence Learning. Nature and Science of Sleep, 16, 879–896. 10.2147/NSS.S463495

Zhang, G.-Q., Cui, L., Mueller, R., Tao, S., Kim, M., Rueschman, M., Mariani, S., Mobley, D., & Redline, S. (2018). The National Sleep Research Resource: Towards a sleep data commons. Journal of the American Medical Informatics Association, 25(10), 1351–1358. 10.1093/jamia/ocy064

